# A Phase I, Randomized, Double-Blind, Placebo-Controlled Trial of Intracisternal Engineered Autologous Exosomes (NV-101) to Restore Neuronal Proteostasis in Early Alzheimer’s Disease: The RESTORE Protocol

**DOI:** 10.1101/2025.09.04.25335118

**Authors:** Minahil Iqbal

**Affiliations:** Iqra University

## Abstract

**Background:** The current therapeutic landscape for Alzheimer’s disease (AD) offers limited efficacy. The failure of the neuronal proteostasis network a system critical for preventing toxic protein aggregation is a fundamental pathological process in AD. Molecular chaperones, which are deficient in the AD brain, represent a compelling therapeutic target. A major obstacle has been delivering large biologics like chaperones across the blood-brain barrier. Engineered exosomes offer a promising platform for neuron-specific delivery of nucleic acid cargo.

**Objective:** The primary objective is to evaluate the safety and tolerability of a single intracisternal dose of NV-101, an autologous dendritic cell-derived exosome engineered with neuron-targeting peptides and DNAJB6 mRNA, compared to placebo in participants with early AD. Secondary objectives are to assess its biological activity through cerebrospinal fluid (CSF) biomarkers.

**Methods:** The RESTORE trial is a single-center, randomized, double-blind, placebo-controlled, dose-escalation study. Eighteen participants with biomarker-confirmed early AD will be enrolled across three sequential dose cohorts (Low, Medium, High). Participants will be randomized in a 2:1 ratio to receive either NV-101 or placebo (excipient buffer) via a single intracisternal magna injection. The primary outcome is the incidence of treatment-emergent adverse events over 52 weeks. Secondary outcomes include change from baseline in CSF levels of DNAJB6 protein, oligomeric amyloid-β42, and phosphorylated tau (p-tau181). Exploratory outcomes include neuroimaging and cognitive measures.

**Conclusions:** This first-in-human study will generate critical safety and initial proof-of-mechanism data for a novel therapeutic strategy aimed at correcting proteostatic failure in AD.

## 1. Introduction & Rationale

Alzheimer’s disease remains the leading cause of dementia worldwide. The recent development of anti-amyloid immunotherapies, such as lecanemab and donanemab, represents a significant advance; however, their clinical benefits are modest and are associated with substantial safety risks, including amyloid-related imaging abnormalities (ARIA) [1, 2]. These limitations underscore that amyloid clearance alone, particularly after clinical onset, may be insufficient to halt disease progression, emphasizing the urgent need to target earlier, more fundamental pathological drivers.

A pivotal upstream mechanism is the collapse of the neuronal proteostasis network. This sophisticated system, comprising molecular chaperones, the ubiquitin-proteasome system, and autophagy, is essential for maintaining protein homeostasis by ensuring proper folding and clearance of misfolded proteins. An age-related decline in proteostasis capacity is hypothesized to create a permissive environment for the aggregation of amyloid-β (Aβ) and tau [3]. The DNAJ/HSP70 chaperone system is a crucial component of this network. Specifically, the DNAJB6 chaperone has been demonstrated to be a potent suppressor of amyloid fibril formation in models of protein aggregation, such as polyglutamine diseases [4]. A broader deficiency in chaperone function and proteostatic regulation is a recognized hallmark of aging and neurodegenerative pathologies [5], making the restoration of chaperone activity a rational therapeutic strategy for AD.

The principal challenge in translating this approach has been the targeted delivery of a large protein-based therapeutic, such as a chaperone, across the blood-brain barrier (BBB). Systemically administered biologics typically exhibit negligible brain penetration. Exosomes, naturally occurring extracellular vesicles, have emerged as an ideal delivery vector. They are inherently biocompatible, can be loaded with various therapeutic cargos (including proteins and RNAs), and can be engineered for enhanced tissue targeting. Foundational work by Alvarez-Erviti et al. showed that exosomes engineered to express the neuron-targeting peptide RVG on their surface could deliver siRNA specifically to neurons in the mouse brain following systemic injection, resulting in functional gene knockdown [6]. This breakthrough established exosomes as a powerful and versatile platform for CNS drug delivery.

The RESTORE protocol synthesizes these two advanced scientific concepts. We hypothesize that intracisternal administration of autologous exosomes, engineered with RVG-Lamp2b for neuronal targeting and loaded with DNAJB6 mRNA (NV-101), will be safe and well-tolerated. We further hypothesize that this intervention will lead to neuronal transduction, increased expression of DNAJB6 protein, and a consequent reduction in the concentration of pathogenic soluble oligomers of Aβ and tau in the CSF of individuals with early AD. This first-in-human trial is designed to test the foundational safety and mechanistic premises of this innovative approach.

## 2. Objectives

### 2.1. Primary Objective

To determine the safety and tolerability of a single intracisternal dose of NV-101 compared to placebo in patients with early Alzheimer’s disease over a 52-week observation period.

### 2.2. Secondary Objectives

To characterize the pharmacokinetic and pharmacodynamic profile of NV-101 by assessing:

**a. Pharmacokinetics:** The change from baseline in CSF concentration of human DNAJB6 protein.

**b. Pharmacodynamics:** The change from baseline in CSF concentrations of:

i. Oligomeric Amyloid-β42 (oAβ42)
ii. Phosphorylated tau (p-tau181)

### 2.3. Exploratory Objectives

To explore the effects of NV-101 on:

a. Brain amyloid burden measured by [[18]F]florbetaben PET and tau burden measured by [[18]F]flortaucipir PET at Week 52.
b. Cognitive and clinical function assessed by the CDR-Sum of Boxes (CDR-SB), Alzheimer’s Disease Assessment Scale–Cognitive Subscale (ADAS-Cog14), and Mini-Mental State Examination (MMSE) at Weeks 12, 24, and 52.
c. Biodistribution of a co-administered dose of superparamagnetic iron oxide nanoparticle (SPION)-labeled NV-101 exosomes via magnetic particle imaging (MPI) at 24 and 72 hours post-injection.

## 3. Study Design

This is a single-center, randomized, double-blind, placebo-controlled, dose-escalation study. The trial will utilize a classic 3+3 design for safety monitoring during dose escalation. Three sequential dose cohorts are planned (Low, Medium, High). Within each cohort, six participants will be randomized in a 2:1 ratio to receive either NV-101 or placebo. All study participants, their care partners, investigators, outcome assessors, and site staff (except for the unblinded Investigational Drug Pharmacist) will be blinded to treatment assignment. The total study duration for each participant will be approximately 60 weeks, including a screening period, a 4-6 week period for leukapheresis and NV-101 manufacturing, and a 52-week post-treatment observation period.

## 4. Methods

### 4.1. Participant Selection

#### Inclusion Criteria

Participants must meet all of the following criteria:

1. Aged 55 to 80 years, inclusive.

2. Diagnosis of mild cognitive impairment or mild dementia due to Alzheimer’s disease consistent with NIA-AA criteria [7].

3. Biomarker confirmation of AD pathology, defined by a CSF profile (low Aβ42/40 ratio and elevated p-tau181) or a positive amyloid-PET scan within the past 12 months.

4. Clinical Dementia Rating (CDR) global score of 0.5 or 1.0.

5. Stable dose of permitted AD medications (e.g., acetylcholinesterase inhibitors) for ≥8 weeks prior to screening.

6. Has a reliable study partner who can accompany the participant to visits.

7. Provides written informed consent.

#### Exclusion Criteria

Participants meeting any of the following criteria will be excluded:

1. Significant neurological disease other than AD (e.g., Parkinson’s disease, significant stroke, epilepsy).

2. Major psychiatric disorder (e.g., major depressive episode, schizophrenia) within the past year.

3. Contraindications to MRI, lumbar puncture, general anesthesia, or intracisternal injection.

4. History of immunosuppression or current use of immunomodulatory drugs.

5. Abnormal laboratory values indicative of clinically significant uncontrolled hematologic, renal, hepatic, cardiac, or infectious disease.

6. Participation in another investigational drug trial within 60 days or 5 half-lives prior to screening.

### 4.2. Randomization and Blinding

Randomization will be performed by the institution’s Investigational Drug Pharmacy using a computer-generated scheme stratified by dose cohort. The pharmacy will be responsible for labeling the sterile study product syringes with identical labels containing only the participant ID and protocol number. The placebo is a sterile buffer (PBS with 10% sucrose) that is visually indistinguishable from the NV-101 formulation. The blinding will be maintained for the entire study duration unless unblinding is required for urgent medical management, as determined by the Data and Safety Monitoring Board (DSMB).

### 4.3. Intervention: NV-101 Manufacturing and Control

#### 4.3.1. Leukapheresis & Dendritic Cell Generation

Eligible participants will undergo a single leukapheresis procedure to collect peripheral blood mononuclear cells (PBMCs). Monocytes will be isolated by adherence or magnetic selection and differentiated into immature dendritic cells (iDCs) over 7 days using recombinant human GM-CSF (800 IU/mL) and IL-4 (500 IU/mL) in serum-free media, as described in established protocols [8].

### 4.3.2. Exosome Isolation and Engineering

Exosomes will be harvested from iDC culture supernatants by sequential ultracentrifugation (300 ×g for 10 min, 2000 ×g for 20 min, 10,000 ×g for 30 min, and 120,000 ×g for 70 min). The pellet will be resuspended in sterile PBS. Exosomes will be characterized for size, concentration, and marker expression.

### 4.3.3. Critical Quality Attributes (CQAs)

Each batch of NV-101 must meet the following pre-defined release criteria before administration:

▪ **Size & Concentration:** 80-150 nm diameter, concentration ≥ 5×10^10 particles/mL (via Nanoparticle Tracking Analysis).
▪ **Purity:** Particle-to-protein ratio > 3×10^10 particles/μg (BCA assay).
▪ **Surface Markers:** Positive for exosomal markers CD63 and CD81 (>90% by flow cytometry); Positive for RVG-Lamp2b (>50% by flow cytometry).
▪ **mRNA Payload:** Loading of ≥ 1000 molecules of DNAJB6 modRNA per exosome (via qRT-PCR).
▪ **Sterility:** Negative bacterial and fungal culture for 14 days.
▪ **Endotoxin:** <0.5 Endotoxin Units (EU)/mL (Limulus Amebocyte Lysate test).
▪ **Mycoplasma:** Negative.

### 4.3.4. Electroporation

Isolated exosomes will be electroporated (using a Bio-Rad Gene Pulser Xcell system) with nucleotide-modified DNAJB6 mRNA (modRNA) using optimized parameters (0.4 mg/mL RNA, 500 V, 125 μF, 200 Ω) to ensure efficient loading without vesicle aggregation, based on principles of mRNA loading [9].

#### 4.3.5. Placebo

The placebo control is the final formulation buffer (PBS with 10% sucrose) processed through an identical mock manufacturing run, including passage through the electroporation system without exosomes or mRNA.

### 4.4. Administration

The intracisternal magna injection will be performed by a qualified interventional neuroradiologist under general anesthesia in an angiography suite. Using strict aseptic technique and real-time fluoroscopic guidance, a 22-gauge spinal needle will be advanced into the cisterna magna. Correct placement will be confirmed by the free flow of CSF. The study product (NV-101 or placebo) will be injected slowly over 5 minutes. Participants will be transferred to the neuro-intensive care unit for a minimum of 48 hours of continuous neurological monitoring for potential adverse events.

### 4.5. Outcome Measures

#### 4.5.1. Primary Outcome

▪ **Safety and Tolerability:** Incidence, severity, and relatedness of all Treatment-Emergent Adverse Events (TEAEs) and Serious Adverse Events (SAEs), with specific attention to predefined Adverse Events of Special Interest (AESIs): aseptic meningitis, intracerebral hemorrhage, seizures, new cranial neuropathies, and significant neurological deterioration.

#### 4.5.2. Secondary Outcomes

▪ Change from baseline in CSF concentrations at Weeks 1, 4, 12, 24, and 52 for:
✓ DNAJB6 protein (quantified by ELISA)
✓ Oligomeric Amyloid-β42 (oAβ42) (quantified by MSD or Simoa immunoassay)
✓ Phosphorylated tau (p-tau181) (quantified by MSD or Simoa immunoassay)

### 4.5.3. Exploratory Outcomes

▪ Change from baseline in brain amyloid burden ([[18]F]florbetaben PET) and tau burden ([[18]F]flortaucipir PET) at Week 52.
▪ Change from baseline in cognitive scores: CDR-Sum of Boxes (CDR-SB), Alzheimer’s Disease Assessment Scale–Cognitive Subscale (ADAS-Cog14), and Mini-Mental State Examination (MMSE) at Weeks 12, 24, and 52.
▪ Biodistribution of a subset of SPION-labeled NV-101 exosomes via MRI/MPI at 24 and 72 hours post-injection.

### 4.6. Safety Monitoring

An independent Data Safety Monitoring Board (DSMB) will be convened, comprising a neurologist, a neuroradiologist, and a biostatistician. The DSMB charter will include predefined stopping rules (e.g., halting enrollment if ≥2 participants in a cohort experience a Dose-Limiting Toxicity (DLT), defined as any related Grade ≥3 AE or any related SAE). The DSMB will review unblinded safety data after all participants in a cohort complete the Week 4 visit and at predetermined intervals thereafter.

### 4.7. Statistical Analysis

The sample size (n=18) is based on feasibility for a first-in-human study and is sufficient to characterize the safety profile using a 3+3 dose-escalation design. Safety analyses will be descriptive, presenting the number and percentage of participants experiencing TEAEs, SAEs, and AESIs. For biomarker data, longitudinal changes will be analyzed using linear mixed-effects models with time, treatment group, and the time-by-treatment interaction as fixed effects, and participant as a random effect. Given the exploratory and hypothesis-generating nature of this Phase I trial, analyses of biomarker and cognitive data will be interpreted with caution, without adjustment for multiple comparisons. All analyses will be conducted using the intention-to-treat principle.

## 5. Ethics and Dissemination

This protocol, informed consent documents, and Investigator’s Brochure will be submitted for review to the appropriate Institutional Review Board (IRB)/Ethics Committee (EC) and to the relevant national regulatory authority. The trial will be conducted in accordance with the principles of the Declaration of Helsinki and the International Council for Harmonisation Good Clinical Practice (ICH-GCP) guidelines.

Results of this trial, whether positive, negative, or inconclusive, will be disseminated through publication in a peer-reviewed international journal and presentation at major scientific conferences. A de-identified dataset and the statistical analysis plan will be made available to qualified researchers upon reasonable request after the primary publication, to promote transparency and scientific collaboration.

## Data Availability

This protocol will be submitted to ClinicalTrials.gov prior to participant enrollment.

